# Wellbeing Impact Study of High-Speed 2 (WISH2): Protocol for a mixed-methods examination of the impact of major transport infrastructure development on mental health and wellbeing

**DOI:** 10.1101/2024.01.31.24302086

**Authors:** Katherine I. Morley, Lucy Hocking, Catherine L. Saunders, Jennifer W. Bousfield, Jennifer Bostock, James Brimicombe, Thomas Burgoine, Jessica Dawney, Joanna Hofman, Daniel Lee, Roger Mackett, William Phillips, Jon Sussex, Stephen Morris

**Author notes:** Corresponding author (Katherine Morley). Katherine I. Morley* *Roles:* Conceptualization, Data Curation, Funding Acquisition, Investigation, Methodology, Supervision, Visualization, Writing - Original Draft Preparation, Writing - Review & Editing Lucy Hocking *Roles:* Investigation, Project Administration, Resources, Writing - Original Draft Preparation, Writing - Review & Editing Catherine L. Saunders *Roles:* Conceptualization, Data Curation, Formal Analysis, Funding Acquisition, Investigation, Methodology, Supervision, Visualization, Writing - Original Draft Preparation, Writing - Review & Editing Jennifer W. Bousfield *Roles:* Conceptualization, Investigation, Methodology, Writing - Review & Editing Jennifer Bostock *Roles: Public involvement and ethics, reviewing and drafting* James Brimicombe *Roles:* Data Curation, Investigation, Software, Resources, Project Administration Thomas Burgoine *Roles:* Conceptualization, Formal Analysis, Funding Acquisition, Investigation, Methodology, Resources, Writing - Original Draft Preparation, Writing - Review & editing Jessica Dawney *Roles:* Investigation, Project Administration, Writing - Reviewing & Editing Joanna Hofman *Roles:* Investigation, Methodology, Writing - Review & Editing Daniel Lee *Roles:* Data curation, Formal analysis, Software, Visualization, Investigation, and Writing - Review & Editing. Roger Mackett *Roles:* Conceptualization, Writing - Original Draft Preparation, Writing - Review & Editing William Phillips *Roles:* Investigation, Project Administration, Writing - Review & Editing Jon Sussex *Roles:* Conceptualisation, methodology, supervision, writing - review and editing. Stephen Morris *Roles:* Conceptualization, Data Curation, Funding Acquisition, Investigation, Methodology, Supervision, Visualization, Writing - Original Draft Preparation, Writing - Review & Editing. **Funding:** This research is funded by The Department for Transport and High Speed 2 Ltd and is independently managed by the National Institute for Health and Care Research (NIHR132761). The views expressed are those of the authors and not necessarily those of The Department for Transport, High Speed 2 Ltd, or the National Institute for Health and Care Research. **Competing interests**: Roger Mackett is a member of Disabled Persons Transport Advisory Committee which advises the government on the transport needs of disabled people.

## Abstract

Although research has demonstrated that transport infrastructure development can have positive and negative health-related impacts, most of this research has not considered mental health and wellbeing separately from physical health. There is also limited understanding of whether and how any effects might be experienced differently across population groups, whether this differs according to the stage of development (e.g. planning, construction), and how changes to planned infrastructure may affect mental health and wellbeing. This paper presents a protocol for the Wellbeing Impact Study of HS2 (WISH2), which seeks to address these questions using a high-speed rail development in the UK as an applied example. WISH2 is a 10-year, integrated, longitudinal, mixed-methods project using general practices (primary medical care providers in the UK) as an avenue for participant recruitment and for providing a geographically defined population for which aggregated data on mental health indicators are available. The research comprises: (i) a combined longitudinal and repeated cross-sectional cohort study involving multiple waves of survey data collection and data from medical records; (ii) longitudinal, semi-structured interviews and focus groups with residents and community stakeholders from exposed areas; (iii) analysis of administrative data aggregated at the general practice population level; and (iv) health economic analysis of mental health and wellbeing impacts. The study findings will support the development of strategies to reduce negative impacts and/or enhance positive mental health and wellbeing impacts of high-speed rail developments and other large-scale infrastructure projects.

## Introduction

High-speed rail infrastructure has been developed in many countries, such as China, France, Japan, and Germany. Its implementation in the United Kingdom (UK), and elsewhere, has been surrounded by lively debate about the balance between investment costs and likely benefits [1]. A recent meta-analysis found that the majority of studies investigating the impact of high-speed rail system developments have focused on changes in accessibility, modes of transport, the environment, tourism, housing or land, labour market, and economic performance [1]. Very limited consideration has been given to health impacts, particularly impacts on mental health and wellbeing [1,2].

Transport infrastructure, such as railways, can create physical or psychological barriers in the local community that impact health and wellbeing by disrupting social connectedness, reducing opportunities for exercise, and reducing accessibility to local health services and supermarkets with healthy food options [3–6]. Communities can also be cut-off from the surrounding area which can cause problems such as less commercial activity for local businesses [3,4], or poor integration of new and existing transport systems [4].

The potential for differential impacts of transport infrastructure on mental health and wellbeing to contribute to social exclusion is recognised [7]. Impacts may be experienced differently according to life-stage, health status, proximity, employment status, and social support [3,4,8–12]. A 2021 systematic review found mixed evidence of the wellbeing impacts of urban design interventions in vulnerable groups. While some infrastructure changes (e.g. green infrastructure, improved walking/cycle paths and urban regeneration in general) led to improved psychological outcomes for low-income residents and women, others (including motorway development) did not [12].

In addition to limited examination of health impacts, current literature has focused primarily on completed infrastructure projects and their users; there is little research on the impacts of planning and construction stages, or how these are experienced by those who do not use the transport, despite acknowledgement that the social impacts of infrastructure mean local communities are stakeholders regardless of use [4,8,13–17]. Frameworks for measuring the social costs of pre-operational phases of large-scale projects have often been restricted to factors such as pollution, traffic, noise, and safety without explicit consideration of mental health and wellbeing [14,17]. Nieuwenhuijsen & Kreis [17,18] developed a framework focused on differential health impacts of transport infrastructure that explicitly includes mental health and wellbeing consequences. However, it is primarily focused on operational effects and although it considers some pre-operational factors (e.g. land acquisition), not all factors relevant to mental health and wellbeing are captured (e.g. uncertainty over project scale, fears of forced relocation) [16].

Thus, there is a need for more research on the impact of transport infrastructure projects on health, particularly mental health and wellbeing. Although research has demonstrated transport infrastructure can have positive and negative health-related impacts, most of this research has not considered mental health and wellbeing separately from physical health. There is also limited understanding of whether and how any effects might be experienced differently across population groups, whether this differs according to the stage of development (e.g. planning, construction), and how changes to planned infrastructure (e.g. cancellation of parts of a project) may affect mental health and wellbeing. The Wellbeing Impact Study of HS2 (WISH2) seeks to address these questions, using a high-speed rail development in the UK as an applied example.

### Context to the research study

High Speed Two (HS2) is a new high-speed railway being implemented by the UK Government, intended to improve connections between London and major cities in the midlands and north of England. The proposal for HS2 was originally put forward in 2009 [19], with a Y-shaped route connecting London, Birmingham, Manchester and Leeds proposed in 2010 [20] (**Fig 1**). Splitting the route into two phases was first proposed in 2011, with Phase 1 running from London to Birmingham and Phase 2 running from Birmingham to Manchester and Leeds [21]. Phase 2 was then split into two parts – Phase 2a, running from the West Midlands (Birmingham) to Crewe, and Phase 2b involving the route from Crewe to Manchester (“Western leg”), and Birmingham to Leeds (“Eastern leg”) [22]. This research project is focused on the Phase 2 sections of the route.

**Fig 1.**
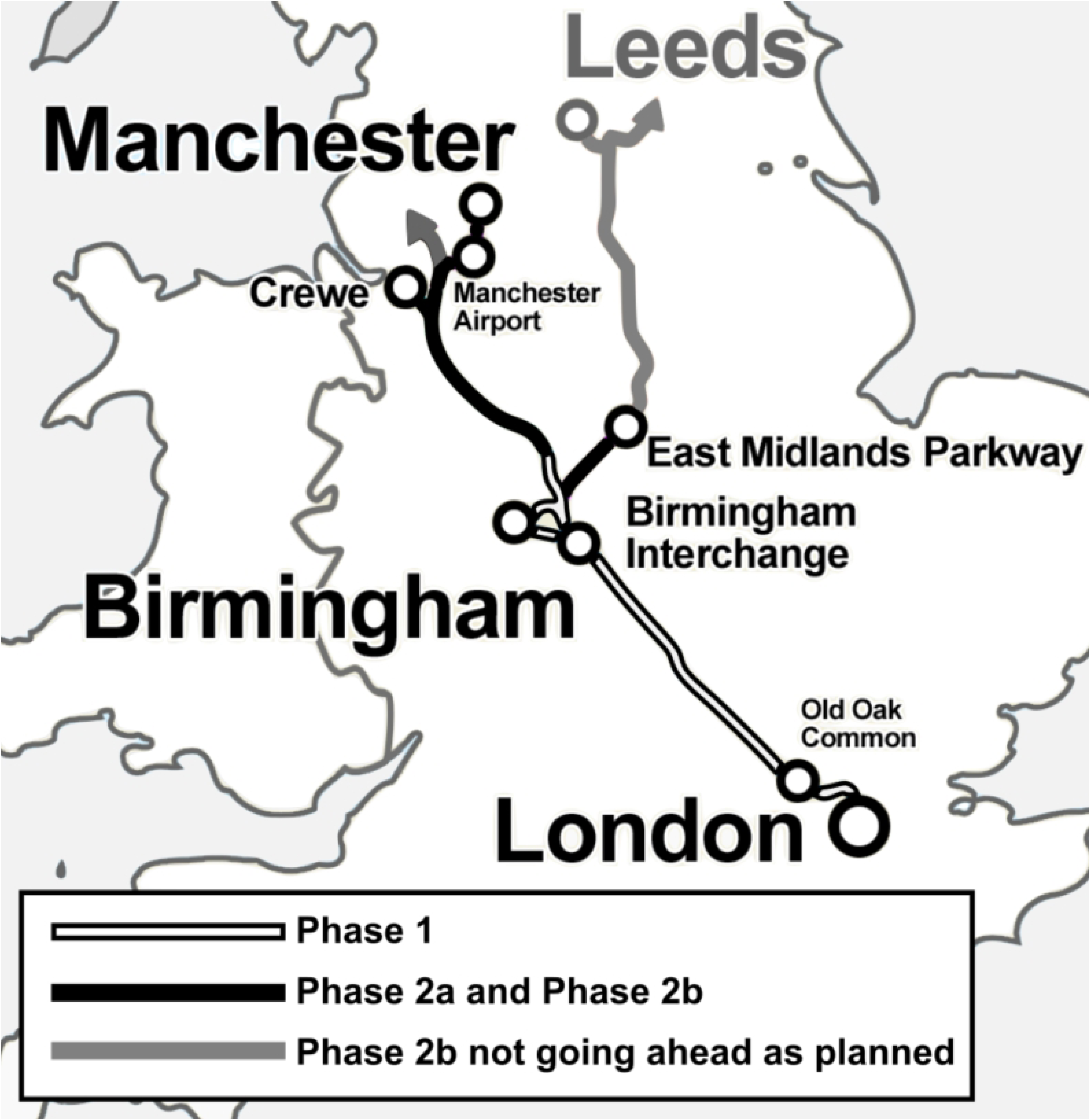
Simplified diagram of HS2 route. Image credit: adapted from image by User:Cnbrb, CC BY-SA 3.0, via Wikimedia Commons.

The necessary legislation to proceed with HS2 was introduced in three parts, aligned to Phase 1, Phase 2a, and Phase 2b sections of the route (see **Fig 2**). In 2013 the UK Government published the High Speed Rail (London-West Midlands) Bill; it received Royal Assent in 2017 [23]. Construction on Phase 1 officially began in 2020, although enabling works at Euston station in London began in 2019 [22].

**Fig 2.**
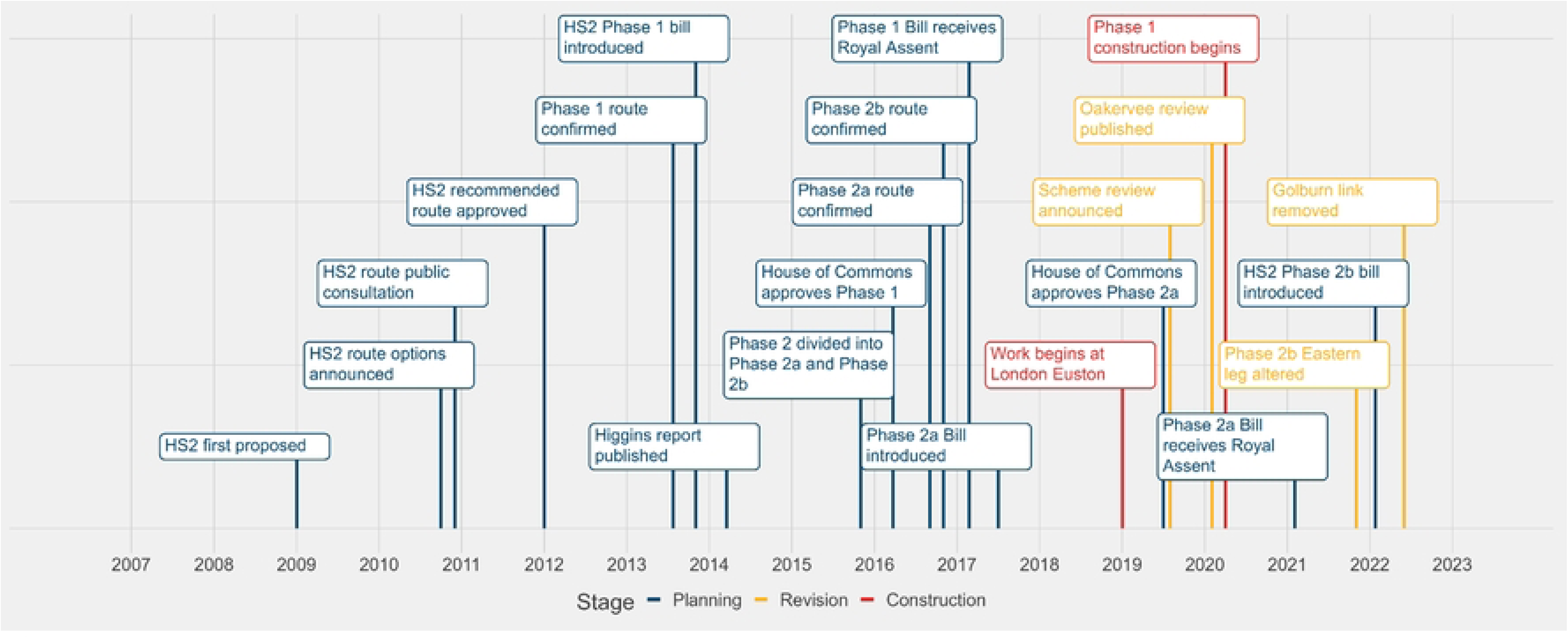
Timeline of major milestones for High-Speed Two development.

The High Speed Rail (West Midlands-Crewe) Bill, covering the Phase 2a section of the route, was introduced in 2017 and received Royal Assent in February 2021 [24]. While some initial work started on Phase 2a following Royal Assent, at the time of writing (mid-2023) construction of the Phase 2a route has been paused until the end of the UK government’s current public spending review period in March 2025 [25]. In November 2021, while Phase 2b was still in the planning stage and six months after this research project commenced, the UK government announced that the HS2 Phase 2b route would not proceed as originally planned. The eastern leg of the Phase 2b route, from East Midlands Parkway to Leeds, and a small section of the route between Warrington and Manchester, will not go ahead as part of HS2. The eastern leg of the Phase 2b route has been replaced with the Integrated Rail Plan (IRP) [26]. The IRP commits to building the Eastern Leg to the East Midlands to serve Nottingham, Derby and Sheffield. The IRP also commits Government to spending £100m on work to look at how best to take HS2 trains to Leeds. While the impact of the IRP is not specifically within the scope of this research, the change of plans means there is still uncertainty for residents and communities on the Eastern Leg of the Phase 2b HS2 following the IRP announcement. The Bill for Phase 2b West, High Speed Rail (Crewe-Manchester) Bill was published in January 2022 and is currently at the Committee stage [27]. No bill for the eastern leg of HS2 Phase 2b has been published as yet.

The Bills for HS2 are hybrid bills, which means that in addition to the standard parliamentary processes, individuals and businesses who are affected by the Bill and its Additional Provisions are able to petition Parliament and the Committee reviewing the Bill [22]. There were many petitions regarding the High Speed Rail (London-West Midlands) Bill relating to effects the HS2 development was having, or was predicted to have, on the mental health and wellbeing of people living along the route [28]. The impact of HS2 on mental health and wellbeing was also highlighted by the House of Commons Select Committee for the High Speed Rail (West Midlands-Crewe) Bill [29]. The Committee directed HS2 Ltd to commission epidemiological research on the impact of HS2 on community mental health and wellbeing. This paper describes the protocol for the commissioned research study, planned to take place over a 10-year period, starting in mid-2021, accounting for the revisions to the plans that were announced in November 2021. This protocol will be amended if further changes to the project are needed as a result of government policy decisions.

### Design

#### Aim

The overarching aim of this research is to investigate the extent to which individuals and communities exposed to the planning, construction and operation of HS2 experience positive or negative mental health and wellbeing impacts, focused specifically on anxiety, depression, and general wellbeing. It will address the following research questions:

1. What are the positive and negative mental health and wellbeing impacts of HS2?
2. Do these impacts change over time and what explains them?
3. Are impacts felt differently across groups within a community?
4. What are the health economic implications of the mental health and wellbeing impacts of HS2?

Due to changes to the HS2 development plan, particularly regarding the Phase 2b East route, the scope of the project was changed during the planning stage to encompass the impacts that may occur when changes are made late in the planning stage. This means that the geographical scope of the study is based on the planned HS2 route as of June 2021 i.e. all parts of the Phase 2a and Phase 2b route shown in **Fig 1**, even those no longer going ahead as planned.

#### Overview and framework

Government policy implementation is usually considered a complex intervention due to the number of components involved, intervention variation over time, and the range of population subgroups affected [30]. The updated Medical Research Council guidance emphasises the importance of mixed methods approaches to intervention research in public health settings [30]. Consequently, for this research we have designed an integrated longitudinal, mixed-methods approach, using general practices as an avenue for participant recruitment and for providing a geographically defined population for which aggregated data on mental health indicators are available. The core data collection elements of the research study are:

- Combined longitudinal and repeated cross-sectional cohort study involving multiple waves of survey data collection and data from medical records (cohort data)
- Semi-structured interviews and focus groups with residents from exposed areas and community stakeholders using the same combined longitudinal and repeated cross-sectional design as the cohort data (qualitative data)
- Analysis of administrative data aggregated at the practice population level (administrative data)
- Health economic analysis of mental health and wellbeing impacts (health economics).

Three waves of data collection and analysis will be undertaken over the 10-year project. These three data collection periods were originally designed to align to the planning, construction, and operation phases of HS2, but this may change if the timeline for the HS2 development is altered. How these data collection activities map back to the research questions is shown in **Table 1**.

**Table 1.** Mapping of research questions to research elements.

| Research Question | Study elements that will address the research question |  |  |  |
| --- | --- | --- | --- | --- |
|  | Cohort data | Qualitative data | Administrative data | Health economics |
| RQ1: What are the positive and negative MHW impacts of HS2? | ✓ | ✓ | ✓ |  |
| RQ2: Do these impacts change over time and what explains them? | ✓ | ✓ | ✓ |  |
| RQ3: Are impacts felt differently across groups within a community? | ✓ | ✓ |  |  |
| RQ4: What are the health economic implications of the MHW impacts of HS2? | ✓ | ✓ | ✓ | ✓ |

To ensure these elements of the research are integrated from the design stage onwards, we developed a logic model to characterise the outcomes and impacts of the HS2 development and develop a framework for all data collection components. We drew on previous transport frameworks and incorporated concepts from the Consolidated Framework for Implementation Research (CFIR) [31–33]. The CFIR assesses implementation barriers, obstacles, enablers, and facilitators with consideration explicitly given to the context(s) of an intervention [33–36]. The logic model combines a conventional logic model with the CFIR, describing how the implementation determinants, implementation strategies and the mechanisms of action they engender, and implementation and clinical outcomes are inter-related [37]. It provides a framework for structured consideration of multiple “context-mechanism-outcome” scenarios as recommended by current guidance [30,38,39], and can be extended to consider both short- and long-term outcomes or impacts.

We developed an initial logic model based on reviewing the academic and grey literature and holding a workshop with people living along the HS2 route. The logic model was refined further through an internal team workshop and review and feedback from the WISH2 Public Advisory Group (PAG: see next section). The current version of the logic model is shown in **Fig 3**, mapping out activities, outputs, outcomes and impacts for the three stages of the HS2 Phase 2 development, as well as modifiers and contextual factors. It articulates, from a theoretical perspective, how HS2 may impact mental health and wellbeing. Below, we briefly explain the current model:

- **Outer setting**: This describes wider context within which HS2 Phase 2 is being implemented, including political, economic and environmental situations and challenges. These include major societal events, such as COVID-19.
- **Inner setting:** This relates to the local area or neighbourhood. It covers high-level descriptions of a region including geographical characteristics, services, and the local economy, and communications between HS2 Ltd and individuals and communities.
- **Individual domain:** This defines and describes the people involved in decision-making and implementation of HS2 Phase 2, as well as the deliverers and recipients of it.
- **Process:** This covers the activities, outputs and outcomes required to deliver the HS2 Phase 2 development. The process of implementing HS2 spans three stages - planning or changes to plans, construction, and operation – each of which may affect mental health and wellbeing via different mechanisms. This central section of the diagram can be read from top-to-bottom (moving through the stages of the development) and left-to-right (moving through activities, outputs and outcomes for each development stage).
- **Modifiers:** *Intrinsic and extrinsic modifiers* influence the extent to which outcomes lead to impacts. These may relate to an individual’s demographic or psychological characteristics and traits (intrinsic modifier(s)) or the profile of the local area an individual lives in (extrinsic modifier(s)). *Intervention modifiers* relate to the ways in which HS2 is implemented as this may affect the extent to which HS2 activities impact mental health and wellbeing.
- **Impacts:** This domain encompasses potential impacts related to mental health and wellbeing at the individual and at the community level. The constructs included are distinct but overlapping and inter-related. There is an anticipated secondary impact of lowered mental health or wellbeing on health and mental health service utilisation.

**Fig 3.**
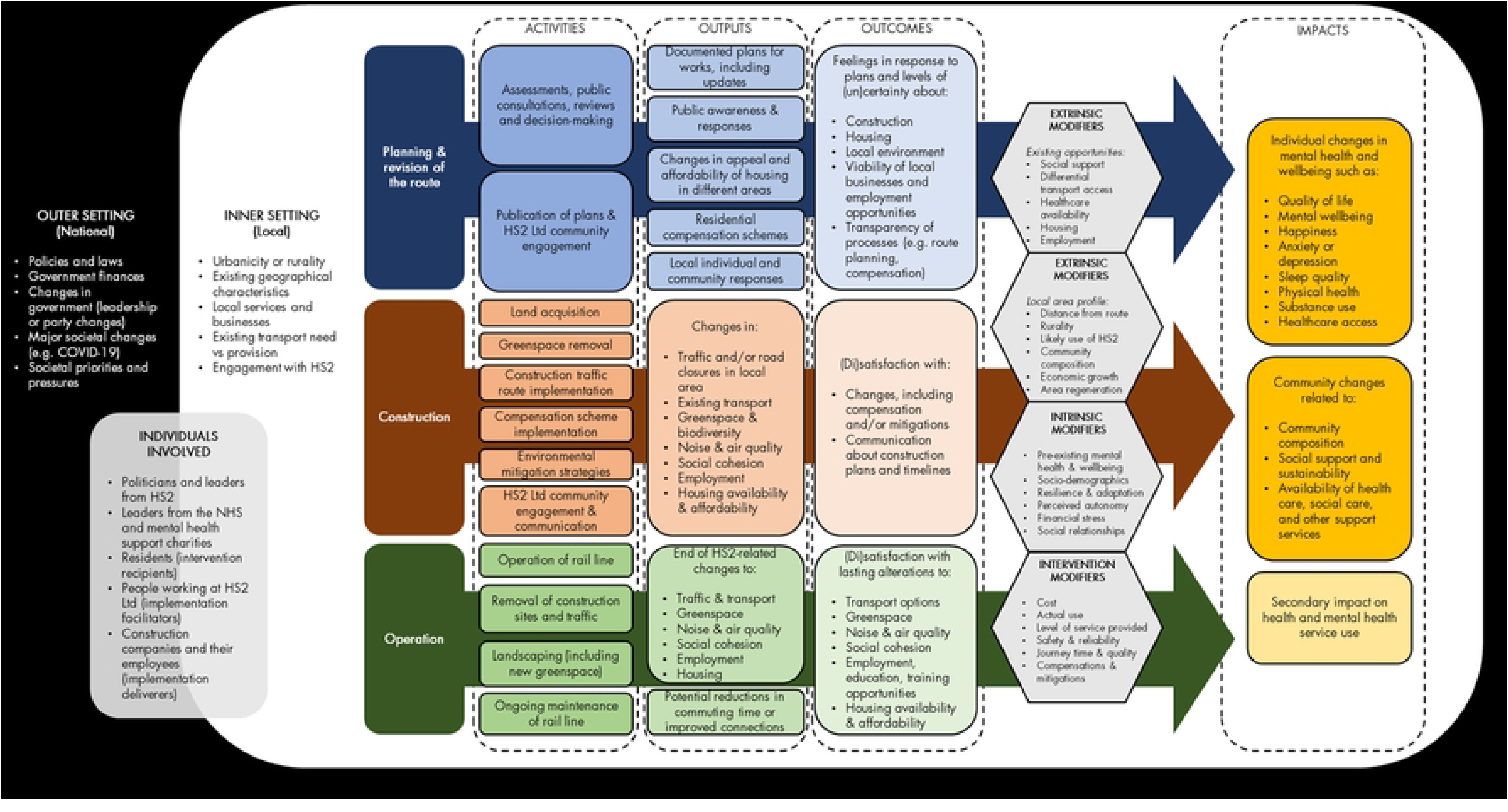
Logic model for HS2 development.

Domains interact with each other, particularly the inner and outer settings. The model does not specify the route of activities through to impacts but this will be developed through the research.

This version of the logic model was used as a framework for the development of the cohort survey, interview and focus group guides for the qualitative research, and the selection of outcomes for the administrative data research for the first wave of the study. Once data are collected, we will use the logic model and underlying CFIR constructs to guide analysis and interpretation. In turn, these findings will be used to update the logic model. We anticipate that as the HS2 development progresses, we will need to develop a separate logic model for the sections of the route that are no longer proceeding as planned due to divergence in relevant activities, outputs, outcomes and impacts.

#### Public involvement

Public involvement is a key part of this study. The research team includes a patient and public involvement Lead who lives on the route of HS1 (Jennifer Bostock) as well as a lay advisor who lives near the HS2 Phase 1 route. The public and patient involvement Lead and lay advisor have convened a Public Advisory Group with six representatives from the general public, including from local communities near the HS2 route. The members are diverse in age, socio-economic status and health and disability status. All members are independent of both the project team and have no links to the transport industry or government. However, some have direct exposure to the construction route. The Public Advisory Group (Lead, lay advisor, and six representatives) is involved in study design, analysis and interpretation and dissemination. The Public Advisory Group meetings are planned for approximately every six months of the study, although meetings may occur more frequently if needed to support the research. This group has already significantly contributed to the design of the study. For example, the group has reviewed draft data collection instruments, consent forms and participant information sheets, providing feedback on how to improve these, particularly the information conveyed and its readability. The Public Advisory Group will advise on and review any changes made to research tools in future study waves. In order to maximise the impact of the study the Public Advisory Group will aid creative dissemination methods and has already been pivotal in the representations of the study on the project website.

## Methods

### Participants and setting

This research involves three groups of participants:

1. Members of the general population, aged 18 and over and registered with a general practice in England;
2. Clinical staff from general practices (General Medical Practitioners (GPs) and Nursing staff);
3. Representatives of community organisations including local government, public health representatives, charities and other support groups, business groups, environmental organisations, and transport organisations.

Participants in group 1 and group 2 will be recruited via general practices, with clinical staff only recruited from practices that are considered “exposed” to the HS2 development (see next section). Participants in group 3 will be drawn from local organisations based in, or that serve the population of, areas in close geographic proximity to the HS2 development. Where necessary to gain a comprehensive understanding of potential impact, organisations with a regional or national perspective will be recruited.

### Exposure definition

For complex, geographically-defined interventions, identifying regions exposed to the intervention and measuring the starting point and degree of exposure are not straightforward; exposure definition is an area of ongoing research. For this research, we use postcodes to determine the geographical proximity of the residential address of individual participants and of general practices to the HS2 development. Individuals are defined as “exposed” to HS2 if their postcode of residence is within 5km of the HS2 Phase 2 route. General practices are defined as “exposed” to HS2 if their postcode is within 5km of the HS2 Phase 2 route *and* at least 50% of the population within the practice catchment live 5km or less from the HS2 route. This geographical area is substantially larger than the area eligible for the HS2 homeowner compensation scheme (<1km from the route) [40], but was chosen to ensure capture of broader impacts such as changes to greenspace or road infrastructure. Unexposed general practices are those with a postcode more than 5km away from the entire HS2 route (including Phase 1) *and* with no-one within their catchment living 5km or less from the HS2 route (i.e. no patients who would be considered “exposed” to any part of HS2). Unexposed participants are recruited from these unexposed practices. General practices not meeting “exposed” or “unexposed” definitions were excluded (e.g. any practices with a postcode within 5km of HS2 Phase 2 but with less than 50% of patients meeting the exposure criteria are excluded, or practices with a postcode more than 5km away from the route but with any patients on the practice list living 5km or less from the route).

### Outcomes

The outcomes for the data collection elements of this research have been selected to support integration of findings from the different data sources. Integration is supported in two main ways: first, where possible, mental health indicators measured in the cohort surveys and medical record data reflect those available in national administrative data aggregated at the general practice level. Second, findings from earlier data collection activities will directly inform subsequent activities. This will occur within each of the three planned data collection waves, with preliminary results from the survey potentially informing changes to data collection instruments for the qualitative research. It will also occur across data collection waves, as data collection instruments for the second and third waves of the study will be directly informed by the findings from previous waves.

#### Cohort data

The primary outcome will be mental wellbeing as measured by the Warwick-Edinburgh Mental Wellbeing Scale (WEMWBS). The WEMWBS scoring range is 14-70. As it approximates a normal distribution it is designed to be analysed using parametric approaches. The minimally important level of change is between 3 and 8 points [41,42]. We will analyse the following secondary mental health outcomes, most of which will also be included in the analysis of administrative data:

- Anxiety and depression incidence. This will be measured via a review of general practice notes using the general practice Quality and Outcomes Framework (QOF; https://digital.nhs.uk/data-and-information/data-tools-and-services/data-services/general-practice-data-hub/quality-outcomes-framework-qof) Systematized Nomenclature of Medicine (SNOMED) Clinical Terms codelists;
- Use of community-provided psychological therapy services. This will be measured using data from the Improving Access to Psychological Therapies national data set (IAPT; https://digital.nhs.uk/data-and-information/data-collections-and-data-sets/data-sets/improving-access-to-psychological-therapies-data-set);
- Prescribing of antidepressants and anxiolytics. This will be via review of general practice prescribing records;
- Prevalence of long-term mental health problems. This will be measured via a question from the GP Practice Survey (GPPS; https://gp-patient.co.uk/) that will be included in the study survey.

Other potential secondary outcomes include alcohol use, and social support (measured via survey questions). We will also collect health and social care service utilisation data (primary and secondary care, personal social services, medications) for the health economic analysis.

#### Qualitative data

While qualitative research does not define “outcomes” in the same way as quantitative research, the topics included in the guides for the interviews and focus groups are informed by the logic model framework (see Design section), the outcomes used in the cohort and administrative data research, and findings from the survey. In addition to standard focus group and interview approaches we will use an adapted version of the Most Significant Change technique [43,44] to collect stories of change to answer RQ2. This technique is well suited to interventions where anticipating all changes in advance is difficult. The method involves defining domains of change (e.g. quality of life, mental health, wellbeing), collecting significant stories of change and selecting the most significant of these.

#### Administrative data

Mental health will be indexed via four related measures:

- Prevalence of self-reported long term mental health problem (GPPS)
- Incidence of new depression diagnoses in people age 18+ (QOF)
- Prevalence of depression in the GP practice population in people age 18+ (QOF)
- Rate (per 1,000 people in the practice population per month) of antidepressant medication prescriptions (from OpenPrescribing; https://openprescribing.net/)

The main outcome will be prevalence of long-term mental health conditions from GPPS. This measure has the advantage of capturing mental health conditions even if a diagnosis has not been entered in the primary care record (e.g. because of self-referred to IAPT). These data are available at least yearly (although during the COVID-19 pandemic recording of some data, particularly QOF, was suspended).

### Sampling and eligibility

We will use a nested sampling design, recruiting general practices and then recruiting participants from those practice populations. A review of GP patient lists by the Office for National Statistics found them to be a reliable population register [45], making this sampling approach broadly equivalent to a traditional address-based sampling approach.

To define “exposed” and “unexposed” general practices we will use geocoded practice postcodes and practice boundary files to identify practices with addresses within 5km of the route of HS2, and practices with boundaries wholly outside 5km of the proposed route. We will also calculate the percentage of each practice population living within 5km of the route of HS2 using the mid-2020 population estimates for Lower layer Super Output Areas (a administratively-defined geographical area consisting of between 400 and 1,200 households; equivalent to a resident population of 1,000 to 3,000 people )[46]. We will exclude practices with less than 750 registered patients to ensure it is feasible to recruit sufficient participants; this is in line with the criteria the Office for Health Improvement & Disparities applies to determine whether practices are included in public health surveillance data sets (https://fingertips.phe.org.uk/profile/general-practice; accessed 17/06/2022). We will also apply an additional criterion for the “unexposed” practices by aiming to recruit those with similar practice population profiles to the “exposed” practices. For each “exposed” practice we will identify a set of “unexposed” practices with similar practice population characteristics using propensity score matching and aim recruit one of the sets to the study. Within “exposed” or “unexposed” practices, adult patients (aged 18 and over) whose registered residential address falls within the “exposed” or “unexposed” geographical regions, respectively, will be eligible for participation in the cohort data collection.

For the qualitative data collection, GP clinical staff will be recruited from “exposed” practices participating in the study. Other community stakeholders will be identified based on the geographical area served by their organisation and/or its physical proximity to the HS2 route. Local residents will be drawn from cohort participants. The survey data will be used to implement a maximum variation sampling approach to select a diverse set of participants. This approach is suggested for mixed-methods studies where breadth of sampling will complement a quantitative analysis [47]. We will ensure participant diversity on factors including gender, age, ethnicity, employment, education, general practice, distance of their residence from HS2 route, and mental health status.

For the cohort surveys and the qualitative research with local residents, we will aim to engage with the same participants at each of the three planned waves of data collection. However, as this will occur over a 10-year period, we anticipate that there will be loss to follow-up. To mitigate this, we will used a combined longitudinal and repeated cross-sectional approach, which has been used in long-term studies of infrastructure project health impacts to manage participant attrition [48]. This approach is illustrated for two waves in Fig 4. Wave 1 participants will initially be directly invited to participate at subsequent waves, but we will recruit new participants from GP practices to make up for any reduction in cohort size over time.

**Figure 4.**
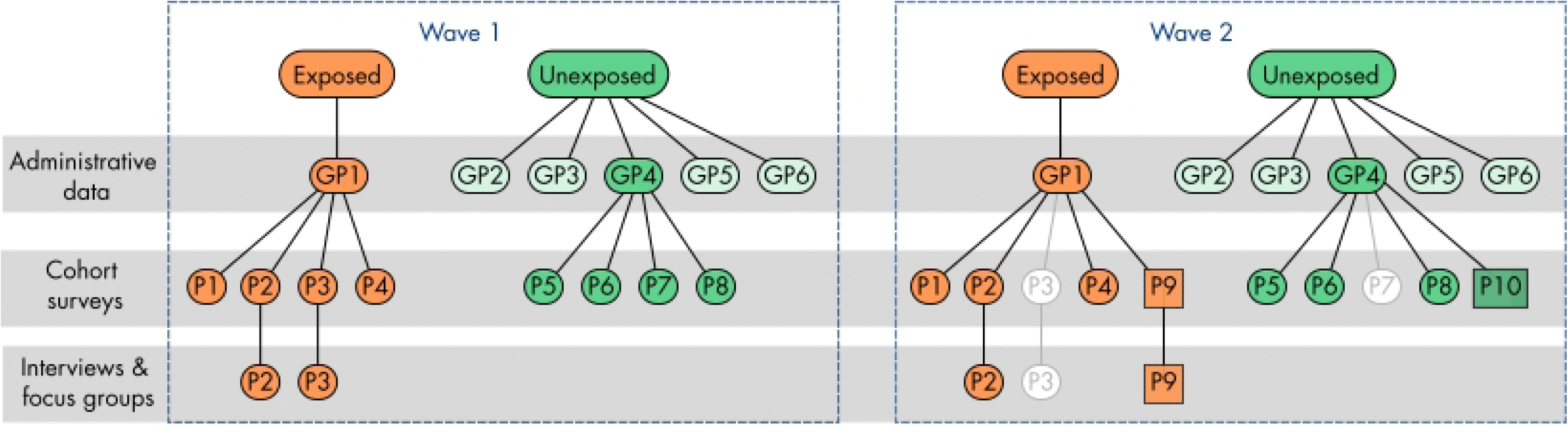
Overview of sampling approach, simplified to illustrate case/control matching of general practice (GP) and patient (P) recruitment for a single exposed practice over two waves of data collection. White circles indicate drop-out at wave 2; squares indicates new patients recruited at wave 2.

### Sample size

#### Cohort data

We aim to survey 6,000 people per wave in four groups (Table 2). General practices through which potential participants will be contacted will be recruited via the National Institute of Health and Care Research Clinical Research Network (CRN).

**Table 2.** Number of GP practices and participants to be recruited per HS2 route segment and by exposure status.

| Route segment |  | Number of general practices | Number of participants per practice (minimum) | Total number of participants |
| --- | --- | --- | --- | --- |
| Phase 2a | Exposed | 12 | 100 | 1,200 |
|  | Unexposed | 12 | 100 | 1,200 |
| Phase 2b West | Exposed | 6 | 100 | 600 |
|  | Unexposed | 6 | 100 | 600 |
| Phase 2b East | Exposed | 6 | 100 | 600 |
|  | Unexposed | 6 | 100 | 600 |
| Phase 2b amended route | Exposed | 6 | 100 | 600 |
|  | Unexposed | 6 | 100 | 600 |
| Grand total |  | 60 | - | 6,000 |

Inclusion criteria are: participants must be age 18 or older and, for “exposed” participants, have a registered address within 5km of the HS2 route. We will ask general practices to exclude patients who are resident in a care home, terminally ill or receiving palliative care, and practices will also have the discretion to exclude patients for other reasons (e.g. because they have requested not to be contacted about anything not directly related to their care, or because they do not have capacity to consent).

#### Qualitative data

For local residents we will conduct eight focus groups per data collection wave, two from each part of the Phase 2 route (Phase 2a Phase 2b West, Phase 2b East, and the section of Phase 2b East that is not proceeding as originally planned), offering one virtual and one in-person focus group to participants from each route section. Each focus group will include a maximum of 10 participants. We will also conduct 3-4 interviews for each route segment. We will conduct up to 53 interviews per data collection wave with clinical staff from participating exposed GP practices and community stakeholder representatives. These sample sizes have been determined in collaboration with the funder and should be sufficient to reach thematic saturation.

#### Administrative data

We will include all general practices in England that have at least 750 registered patients for at least one year during the study period, including those that were active from 2013 onwards. This will result in a total sample of approximately 8,000 practices (in 2013), about 1,000 of which meet the criteria for being “exposed” to the HS2 development. These numbers are approximate because the exact number of practices changes each year as practices close down or merge with others, and new practices open.

### Data collection

#### Cohort data

Potential participants will be contacted via their general practices using a secure mailing service that is commonly used by practices for mailouts. This approach ensures that the research team will not have access to any personal identifying information before individuals have consented to participate in the study. Potential participants will receive a cover letter, participant information sheet, consent form, refusal slip, paper survey booklet, and pre-paid reply envelope in the post. Participants will have the option to complete the refusal slip or consent form and survey using the paper forms or using an online platform (Qualtrics). If preferred, participants will be able to schedule a telephone interview with the research team in which they will go through the survey over the telephone and the staff member enters their responses into the online survey for them. Recruitment for each practice will be open for at least two months, therefore participants have this length of time to decide whether they would like to participate. If required due to low participation rates, one reminder will be sent via text message containing a greeting, a personalised link to the survey.

Survey questions for wave 1 have been selected to reflect the outcomes and potential explanatory factors highlighted in the WISH2 logic model. Where possible, we have used questions from national surveys as this will enable us to compare our data with national estimates. An overview of the topics to be covered and question sources is provided in Table 3.

**Table 3.** Overview of topics covered in the WISH2 survey and sources of questions used.

| Area | Topics covered | Question sources |
| --- | --- | --- |
| Impact of HS2 | Awareness of HS2 development<br>Perspectives on individual and community impact | WISH2 team – developed in collaboration with the PAG |
| Health, wellbeing and healthcare use | Mental wellbeing<br>Physical and mental health conditions<br>Health and social care service use<br>Sleep quality<br>Alcohol use | Warwick-Edinburgh Mental Wellbeing Scale (WEMWBS) [49]<br>EuroQol-5D [50]<br>General Practice Patient Survey [51]<br>Social and Community Opportunities Profile [13,17,18]<br>National Health Sleep Awareness Project [52]<br>Alcohol Use Disorders Test [53] |
| Community and social inclusion | Social networks<br>Financial wellbeing and employment | Social and Community Opportunities Profile [13,17,18] |
| Socio-demographic | Employment<br>Caring responsibilities<br>Transport use<br>Ethnicity<br>Household size | Understanding Society [54]<br>Social and Community Opportunities Profile [13,17,18]<br>NatCen National Transport Survey [55]<br>General Practice Patient Survey [51]<br>Note: Age, gender, and address will be available from GP records so have not been included here to reduce participant burden. |

Consent for access to medical record data (practice data and IAPT data) will be sought from participants at baseline, as well as consent to re-contact for study updates and follow-up. Participants will also be asked to indicate whether they would be willing to participate in interviews or focus groups as part of the study.

#### Qualitative data

Participants selected from the study cohort will be invited to participate in a focus group or interview via telephone, email, or post (for survey participants, based on indicated communication preferences in their survey consent form). Other potential participants (clinical staff and community stakeholders) will be invited by email or telephone. Participants will receive an information sheet and consent form in advance and a list of dates and local venues (for focus groups) or tailored arrangements (for interviews). Each focus group will take up to 120 minutes and interviews up to 60 minutes. In addition to standard focus group and interview approaches, we will use an adapted version of the Most Significant Change technique [43,44]. Focus groups and interviews will be used to collect stories of change. Stories may be recorded with participants’ consent and transcribed verbatim.

Recordings will be deleted once transcripts have been finalised. a combination of direct (interviewee’s experience) and indirect (interviewee relating stories of others’ experiences).

#### Administrative data

Area-level administrative data for general practices will be matched via postcodes within a geographical area defined by the practice catchment using publicly available mapping files [56]. These mapping files are updated regularly by practices, which will enable us to account for changes in practice catchments over time (e.g. due to practices merging or a change in catchment population). All data required are drawn from publicly available sources.

We will use the same sampling approach as described above, but all potential “exposed” and “unexposed” practices will be included, and not just those that are recruiting survey participants. For our initial analyses, general practices in the area of the route that is not proceeding as planned will not be treated separately as the data we are using in the first wave pre-dates the cancellation announcement for the first wave. For subsequent waves, we will treat the general practices from these regions separately from those in areas where the HS2 route is proceeding as planned. We will include a sample of general practices from other areas of England as a second “control” group.

### Analysis plans

We will pre-register an analysis protocol for each wave of data collection, for each data source (cohort surveys, interviews and focus groups, and administrative data) on the Open Science Framework (DOI 10.17605/OSF.IO/HQGDT).

#### Cohort data

To examine the impact of HS2 on outcomes over time, we will use difference-in-differences (DiD) analysis [57]. This quasi-experimental design compares outcomes between groups over time to assess the impact of a policy or environmental intervention. It is an established approach for analysing data from natural experiments where exposure to the intervention is outside control of the researcher, as is the case with the HS2 development [58,59]. The main impact analysis will be conducted separately for Phase 2a, Phase 2b, and the part of the route not proceeding as planned. We will estimate the overall intervention effect and investigate the differences across key subgroups, such as age or housing status, via exploratory subgroup analyses. We will use appropriate univariable and multivariable models, depending on the outcome, and will adjust for baseline characteristics and sampling.

We will explore the impact of exposure in two different ways: area-based and individually computed distance [60]. Our primary analyses will use the area-based definition to enable comparison between individuals living in unexposed areas and individuals living in the exposed areas, which will necessarily treat all individuals as equally exposed. However, as proximity to the HS2 route is likely to be an important factor for mental health and wellbeing outcomes, for our secondary analyses we will focus on those living in exposed areas and define exposure using individually computed distance from the route i.e. creating a gradient of exposure.

We considered the sample sizes needed for 90% power based on a 0.05 significance level for a two-sided test. All calculations were conducted using Stata version 15.0. We examined this for our three main analytical approaches:

1. **Cross-sectional (within-wave) primary outcome analyses:** Data from 2011 estimated the mean WEMWBS for adults in England as 51.6 (standard deviation 8.7) [61,62]. Based on this, a sample size of 178 per group (356 in total for each HS2 Phase) is required to detect a minimally important difference of 3 points in the WEMWBS between intervention (n=178) and control (n=178) groups. Our target sample sizes far exceed this.
2. **Longitudinal (across-wave) primary outcome analyses:** The DiD approach used for the main analysis is a test of an interaction, rather than the test of main effect (simple difference in means); sample size requirements for tests of interactions are four times those required to test a main effect of the same size [63]. Therefore, 356 responses would be needed from both intervention and control cohorts at each of two time points for the same power from this analysis framework. As the recruitment target is 1,200 participants per group per HS2 Phase (n = 4,800 in total), and 600 participants per group for the part of the route not proceeding as planned (1,200 in total) the study will be well-powered to detect a meaningful difference in the primary outcome using a DiD framework, and separate analyses for Phases 2a and 2b will be feasible.
3. **Longitudinal subgroup analyses:** Differential impacts of HS2 on particular groups in the population will be approached analytically as a “triple difference” (difference in difference in difference) analysis (a three-way interaction). It would require a further fourfold increase in sample size for a differential impact of the same size as the main effect meaning an overall sample size of 5,696 would be required (or possibly greater if the groups being compared are not of equal size). Using all three waves of data collection or combining both Phase 2a and Phase 2b sites we would achieve this sample size. However, we will not have sufficient power to undertake subgroup analyses for the section of the route which is not proceeding as planned.

#### Qualitative data

Interview and focus group transcripts will be analysed using MAXQDA. A coding framework developed from the topic guides and WISH2 framework will be used to systematically and deductively code and analyse data. The framework will allow flexibility to capture issues from the data and from the project over time and provide a basis for mapping the evidence from qualitative data analysis against research questions.

To interrogate and interpret the qualitative data from focus groups and interviews we will use thematic analysis. Data from local residents and stakeholders will be analysed separately, so we can explore similarities and differences between the two groups. Thematic analysis categorises data and then examines the relationships and meanings in the categories to identify themes [64,65]. We will conduct a cross-sectional analysis of focus groups and interview transcripts to examine the data across each data set, and the thematic analysis to generate and explore themes or patterns of meaning within these two data sets. This will: (i) enable understanding of what transcripts contain; and (ii) identify recurring themes. We will identify recurring themes that reflect specific patterns or meaning found in the data and categorise these by applying codes to portions of data. These codes will describe a range of factors, attitudes, behaviours, allowing us to explore in more detail if and how HS2 affected people’s health and wellbeing and what factors and strategies could facilitate the process of adapting to change [66].

To select the stories for the Most Significant Change analysis, the researchers will read all the stories elicited from interviews, organise them by theme and select 10 that represent different themes and include impacts at the planning, construction, and operation stages, if available. For each of these stories, an anonymous summary will be written that is no longer than a page in length. A workshop will be held with Public Advisory Group members, with facilitation from the research team, to discuss the stories and between them select the three that they feel represent the most significant changes to mental health and wellbeing and/or the process of adapting to change.

#### Administrative data

The main analysis will use a multivariable regression framework, which will compare exposed and unexposed practice populations and look at the difference in differences over time in the four administrative mental health outcomes. For the outcomes that are proportions or counts, we will disaggregate the practice level measures and use a person level modelling framework (with practice level covariates). Models used will depend on the characteristics of the outcome variables, but we anticipate using mixed or fixed linear, logistic and Poisson models.

We will adjust for age, gender, calendar month and index of multiple deprivation an aggregate measure of the degree to which people living in a particular area of the UK have the income, health, education, housing or other resources required to meet their needs (https://www.gov.uk/government/statistics/english-indices-of-deprivation-2019). We will additionally adjust for pre- and post-intervention trends in the outcomes to explore whether these are differential among exposed and unexposed practices. We will weight the contribution of exposed practices based on the percentage of the practice population that live within 5km of the proposed route. Where appropriate, we will assess the parallel trend assumption of the difference-in-difference approach through visual inspection of plots for each outcome. We will evaluate it analytically by conducting sensitivity analyses using the Omitted Variable Framework by Cinelli & Hazlett for regression models and extended by Landon & Zimmerman for DiD analyses [67,68].

### Health economic analysis

#### Change in costs of health and social care use

In the cohort survey, we will collect participant-level data on health and social care service utilisation (primary and secondary care, personal social services, medications) during the previous three months and analyse this using a difference-in-difference approach. For the economic analysis we will apply unit costs to these utilisation measures from published sources [69–71], and repeat the difference-in-difference approach. This will give the participant-level change in costs of health and care service use over the previous three-month period associated with each wave of the HS2 development (including, separately, the Phase 2b sections not proceeding as planned).

We will repeat these analyses for population subgroups (e.g. based on combinations of socioeconomic status, age, gender and urban/rural status). Selection of specific groups will be informed by the cohort surveys and interview and focus group results. The participant-level change in costs for each subgroup will then be extrapolated pro rata to the actual duration of each wave of the development, and the analysis will be repeated for each wave of the development separately for Phase 2a and 2b. This will result in a dataset of changes in health and social care costs per person for each wave of the HS2 development that varies by population subgroup. We will then multiply these cost changes by the number of people in each population subgroup potentially affected by different phases of the HS2 development based on Office for National Statistics and National Health data, as defined according to our sampling strategy for the cohort surveys, to compute the total change in health and social care costs at the population level associated with the HS2 development by phase and wave. This will require data on the total number of people in each subgroup living within 5km of the route, which will be obtained from the sources used for the administrative data analysis. We will undertake extensive, deterministic sensitivity analyses around our central estimates of these potential impacts, varying the difference-in-differences, unit costs, durations and population sizes.

#### Monetised impacts on quality of life

In the survey, we will collect participant-level data on health related quality of life using EQ-5D-5L [72]. We will use the EQ-5D-5L data to compute utility scores for each participant using the Crosswalk Index Value Calculator [73]. We will repeat the difference-in-difference approach described for the survey data using EQ-5D-5L utility scores as the outcome, which will give the participant-level change in utility scores associated with each wave of the HS2 development.

As above, we will repeat this analysis for population subgroups. The participant-level change in utility scores for each subgroup will then be extrapolated pro rata to the actual duration of each wave of the development to compute the change in quality-adjusted life years (QALYs), resulting in a dataset of QALY changes per person for each wave of the HS2 development that varies by population subgroup. We will then multiply these changes by the number of people in each population subgroup potentially affected by different phases the HS2 development (as above) to compute the total QALY changes at the population level, by phase and wave. These values will be monetised by multiplying the total QALY changes by the prevailing values for a QALY, which currently range from £15,000 per QALY on the basis of the opportunity cost to the NHS to £60,000 per QALY on the basis of willingness-to-pay [74–76]. These will also be subject to extensive sensitivity analyses, varying the DiDs, durations, and population sizes as well as the QALY valuations.

#### Triangulation of results and incorporation of stakeholder feedback

The analysis integration plan for the study is informed and driven by the WISH2 framework and the Pillar Integration Process developed by Johnson, Grove and Clarke [77]. The Pillar Integration Process is a structured mixed methods analysis framework that guides the way in which quantitative and qualitative data are integrated [77]. It enables the systematic visualisation of the analysis process in a way that integrated findings can be traced back to their original sources, enhancing transparency and replicability [77]. Value beyond what is brought by the individual methods comes from the additional thinking and processing of data required to bring together and organise findings into the joint display and populate the ‘pillar building’ column, thus increasing the quality of the integration [77]. This process also encourages researchers to reflect on the limitations of each data source, for instance, the appropriateness of the research questions and comprehensiveness of the research tools.

The logic model and research questions will provide a framework for the integration of findings. We will create a joint display for each research question. Theoretically, joint displays are used to present quantitative and qualitative data from the same ‘case’, which can mean the same participant or similar themes/topics, phenomena or settings [78]. For this evaluation, data will primarily be integrated at the level of themes, with labels added to the joint displays so that data organisation is aligned with the WISH2 framework. We will not integrate data at a participant-level, but to support the exploration of health inequalities we may produce separate joint displays that bring together data across sources (survey, administrative data analyses, interviews, and focus groups) for particular groups. The findings from each wave will be used to update the WISH2 framework iteratively and inform the data collection tools and approaches for the next wave.

After wave 2 and wave 3, we will obtain feedback from local stakeholders and the WISH2 Public Advisory Group, including exploration of any gaps identified. Workshop results will be included with other study data in the final synthesis for each wave. Two workshops per wave (4 in total) will be held with local stakeholders in areas from which participants have been recruited and will act as a forum for stakeholders to provide feedback on study results prior to publication.

### Data management

The study has been pre-registered (ISRCTN58916738; https://www.isrctn.com/ISRCTN58916738).

We will pre-register analysis protocols for each WP and each wave of data collection using the Open Science Framework (https://osf.io/hqgdt).

The paper forms used for consent and collection of quantitative interview data, and all personal identifying information of participants will be securely destroyed 12 months after the completion of the study, once linkage between the quantitative survey data and medical records have been completed and data have been checked to ensure that no reference to the original data collection instruments is necessary.

It is a stipulation of the funding for this project that all the data it generates are made publicly available to other researchers in de-identified form. We will share de-identified versions of the data from the surveys, interviews, and focus groups. In the case of the survey data this will consist of the survey responses, age at survey, gender, and distance from any part of the HS2 route but not name, date of birth, address or NHS number. In the case of interviews and focus groups only the anonymised transcripts of these will be shared, not the recordings to preserve privacy. We will share these data using the UK Data Service (https://ukdataservice.ac.uk/). This means only people registered and approved by the UK Data Service will access to information from this study, and we will know who has been given access and for what reason.

### Ethical considerations and declarations

This study has been reviewed and approved by the East of England (Cambridgeshire and Hertfordshire) Research Ethics Committee (22/EE/0292) and the Health Research Authority (IRAS302856). During the data collection phase of the study, participants will be free to withdraw at any time, without giving a reason. Their sensitive personal data will be destroyed and the participant will not be contacted again by the research team for any future waves of data collection. Data already collected as part of the study will be retained. We are aware that some of our participants may find issues that arise during data collection sensitive and potentially upsetting and hence we are being guided by our Public Advisory Group and our Public and Patient Involvement Lead to advise prior to approvals and amendments being sought.

Any amendments to the study protocol related to survey, qualitative interviews, or focus groups will be submitted to the Health Research Authority and Research Ethics Committee for necessary approvals. Other significant amendments will be recorded in the ISRCTN registry entry for the study (www.isrctn.com/ISRCTN58916738) and/or relevant pre-registrations and analysis plans available on OSF (https://osf.io/hqgdt ).

### Dissemination

We are iteratively developing a dissemination strategy in collaboration with our Public Advisory Group. However, we anticipate that it will include:

- Tools for disseminating results for each wave including: funder reports, academic papers and conference attendance, infographic summaries, plain English leaflets, message-led policy briefings, blogs and podcasts/videos.
- Public Advisory Group involvement may include co-authorship, co-presentation and bespoke dissemination for targeted public audiences.
- A public website that tracks progress and provides access to these dissemination tools (this is already in place and has been refined following Public Advisory Group input - www.wish2.org – and will expand as the research progresses).
- Tailored dissemination approaches for different stakeholder groups using different combinations of dissemination tools.
- After wave 2 and wave 3, workshops to obtain feedback from the stakeholder groups. .

### Publication policy

Due to the large-scale nature of the project, we will publish more detailed protocols for different components of the research on the Open Science Framework as it progresses (https://osf.io/hqgdt/). In addition to this protocol paper, the research team are committed to publishing findings from each wave of the study in Open Access journals.

### Status and timeline

This study is intended to last for 10 years (June 2021 – May 2031) and encompass the planning, construction, and potentially operation of HS2 Phase 2 development. The study will include three data collection waves, each involving an approximately 18-month period of revising and implementing protocols, analysis and reporting. Precise timing depends on HS2 progress, but the study approach has been designed so changes to HS2 timelines can be accommodated. Research commenced in June 2021 and ethical approval was received in January 2023. Recruitment of GP practices started in March 2023 and the first survey packs were mailed to participants on the 18^th^ of May 2023.

### Limitations

As discussed in the Introduction, the planning stage of HS2 began in 2009/2010, while this research began in 2021; this means that the study cannot capture a true “baseline” measure of mental health and wellbeing in communities along the route of HS2. Our analysis of administrative data partially mitigates this, but cannot provide the level of detail on mental health and wellbeing that would have been captured by a survey, interviews, or focus groups.

Defining exposure to the HS2 development is not straightforward. While we set a 5km boundary around the Phase 2 HS2 route to define our exposed group (and this is larger than the HS2 homeowner compensation scheme boundary), it is possible that the impact of HS2 extends beyond this. To mitigate this, the interviews with local stakeholders may offer insight into the impacts of HS2 on a larger geographic area which can be used to inform our overall conclusions from the study. In addition, it is still possible participants that we have defined as “unexposed” may be affected by HS2 through impacts on other aspects of their lives such as commuting or place of work. However, our sampling approach is designed to minimise this risk and reduce contamination between the two groups. Additionally, while we are intending to focus on Phase 2 of the HS2 development, those living at the southern end of the Phase 2 route will also have had some exposure to Phase 1 of the development, which is now at the construction stage. Although Phase 1 is not within the scope of this research project, we will be able to try to explore this potential effect using information on the geographical proximity of participants to both HS2 phases.

Recruiting and retaining study participants is likely to be challenging given the length of time of the study. There is likely to be participant drop-out; to mitigate this, we will recruit new participants at wave 2 and 3 to ensure we maintain a sufficient sample sizegeneral. This approach has been used in long-term studies of infrastructure project health impacts to manage participant attrition [48]. Our use of general practices to recruit people will mean that people who are not registered with a GP will be excluded from the research, although the GP registration in England is almost universal.

HS2 can be an emotive topic and can trigger strong reactions in people; as participants will self-select to take part in the study our sample may be skewed towards residents with stronger views and feelings towards HS2, or those who have been impacted to a greater extent by HS2. During the interviews and focus groups, we will ask participants about positive and negative impacts of HS2, as well as attempt to establish where there has been no notable impact, to ensure we explore all possible outcomes in detail.

Finally, as this is an evaluation of government policy, the “exposure” we are investigating is beyond the control of the research team. Given the timeline for the project, and the changes that have already occurred to date, it is possible that further changes will occur over the project’s intended lifetime. This may ultimately preclude assessment of the operational phase of HS2 as part of this research.

## Discussion

The need for this research study was identified by the UK House of Commons Select Committee, driven by public concern regarding the impact of HS2 on mental health and wellbeing, the lack of evidence on the topic, and the need for community support to mitigate any negative impacts. This will be the only controlled, quasi-experimental study of the mental health and wellbeing impacts of transport infrastructure from planning to completion, filling an evidence gap on HS2 and large-scale infrastructure projects more broadly. The entire HS2 scheme is intended to be completed in 2035-40, so the findings of this study will be directly relevant for at least 20 years. However, the study findings will support development of strategies to reduce negative impacts or enhance positive mental health and wellbeing impacts, which is also relevant to other infrastructure projects.

## Data Availability

No datasets were generated or analysed during the current study. All relevant data from this study will be made available upon study completion.

## Acknowledgements

The research team would like to thank the WISH2 Public Advisory Group and the Study Steering Committee for offering their ongoing time and support in designing and conducting the study.

